# Masking significantly reduces, but does not eliminate COVID-19 infection in a spatial agent-based simulation of a University dormitory floor

**DOI:** 10.1101/2021.09.13.21263458

**Authors:** Roxanne McPeck, Krisztian Magori

**Affiliations:** Eastern Washington University

## Abstract

COVID-ADAPT is a stochastic, discrete-space, agent-based simulation model of airborne infection and public health interventions, capable of modeling SARS-CoV-2 transmission. Using a map of a real university dormitory floor, the model simulates agents moving about and potentially infecting each other. Health interventions tested are vaccination and masking, including whether the solitary initial infectious agent was masked. Universal masking with N95 masks and 100% vaccination of susceptible people resulted in significantly lower prevalence after 3 weeks compared to all other scenarios, but still led to a substantial number of infections. Increased vaccination levels from 52% to 100% by itself did not result in a significant difference in prevalence due to symptomatic and asymptomatic breakthrough infections. These results suggest that vaccination alone is insufficient to stem outbreaks, and the best way to reduce COVID-19 infections is to ensure that all infectious people are masked. However, because asymptomatic infections are common, the only way to ensure this is universal masking, which also reduces prevalence by protecting susceptible individuals from infection. Universal masking is the best way forward, especially under threat of the Delta variant, to keep facilities open and safe for occupants, while minimizing the number of infections.

## Introduction

The emergence of SARS-CoV-2, the virus responsible for COVID-19, has inescapably changed life, business, and education across the world. At the authors’ home university, Eastern Washington University (EWU), a long stretch of online-only instruction and greatly reduced university services helped control spread but introduced much disruption to faculty, students, and staff, not to mention the toll of the virus itself on community and individual health. With rising vaccination rates (currently estimated in Spokane County, the home of EWU, at 52.5% of the 12-and-up population fully vaccinated at the time of writing) [1], the university looked forward to reopening with a vaccine mandate, and, most recently imposed by the Washington State governor, a mask mandate. However, the Delta variant that has overtaken the United States introduces much uncertainty into risk calculations.

Against this backdrop, one of the best tools for those looking to make public health decisions for the university and its population are statistical and mathematical models. With limited data due to the still-nascent rise of this pandemic, simulation models can be particularly helpful in making evidence-based decisions. Although many models for COVID have been developed or adapted in the past year and a half (e.g., [2-7]), there is a lack of models that are both agent-based (modeling individual people) and discrete-space (modeling actual, rather than idealized, spaces), a combination which helps to apply simulation to real-world contexts. One model that fills this gap is COVID-ADAPT.

COVID-ADAPT is a stochastic, discrete-space, agent-based simulation model of airborne infection, developed by Dr. Krisztian Magori in response to the COVID-19 pandemic. Agents, or simulated people, move about a map on a grid, each cell of which represents a 2×2 ft. space in a real building. Their movement is constrained by walls and facilitated by open doors, and for now, they move randomly. These agents have the qualities of being vaccinated or not and masked or not, and can exist in one of four classes (susceptible, exposed, infectious, or recovered), with progression through the classes occurring if an agent starts the model as infectious and recovers, or starts as susceptible but becomes exposed during the simulation.

The model is customizable, including having the capacity to use custom-made maps. In this report, we modeled the second floor of Pearce Hall, a dormitory at EWU. This building was selected by maintenance staff as a particularly illustrative case. It is actually a circular building, but for the purposes of the grid-based model, it has been rendered square. The square footage of rooms and the connections and orientation of rooms relative to other rooms have been retained.

Infectious agents release viral particles that contaminate the space they’re in, such that a contaminated room can still infect a susceptible agent even if the infectious agent has recently left. Particles diffuse to fill a room, but large rooms such as the continuous hallway on Pearce’s second floor are broken into “nodes” that experience diffusion separately from surrounding nodes, representing the limited-in-distance diffusion of most expiratory particles and droplets. Viral particles decay at a constant rate in the model, so rooms that have been vacated by an infectious agent eventually return to a non-contaminated state.

Viral contamination is measured in quanta, a concept of airborne diseases originally developed by Wells & Riley that, somewhat circularly, represents an infectious dose sufficient to cause infection in 63% of those exposed (reviewed in [8]). This has the advantage of being possible to back-calculate from known outbreak data, rather than having to be measured experimentally, as would be, for example, an exact number of virions. In addition, it is a relatively solitary pseudo-unit capable of extrapolation to multiple people, as opposed to infectious doses, which can vary person-to-person with such conditions as individuals’ immune competency and health conditions.

Settings affect how quickly, on average, the stochastic movement of agents through the disease progression occurs once they are exposed, as well as whether an exposure event is sufficient to move an agent into the exposed class, both of which are subject to probability functions. Death is not modeled and there is no isolation or contact tracing, so once exposed, the course is inevitable, but the timing is variable agent to agent and simulation to simulation. Vaccine and mask efficacy settings control how much these interventions mitigate exposure, but with longer exposures, more potential exposure events accumulate, leading to a higher likelihood of becoming exposed.

The trials in this study were designed to test the effects of vaccination and masking in a scenario containing a quantity of agents representing double occupancy of the 20 dorm rooms on the map (40 agents). Vaccination and masking are two of the most promoted yet contentious public health interventions. The simulations in this paper attempt to answer the questions of whether vaccination at current (or the ideal 100%) rates is enough to control the spread of COVID-19, and what additional protective effect, if any, masking may have in those contexts.

## Methods

The second floor of the EWU dorm Pearce Hall was used as the map. Simulations were run for 30,000 minutes in-simulation (approximately three weeks) with 40 agents, one of whom began the simulation as infectious. The initial infectious agent was either masked or not masked but was never vaccinated. Because vaccination status does not change the behavior or quanta emission of an agent once infected in the model, it was not necessary or relevant for this agent due to its starting infectious status. The vaccination and masking status of the 39 susceptible agents are additionally manipulated (Table 1). The total prevalence at the end of each simulation (total number in the infectious and recovered classes, representing the total disease burden in that simulation) was recorded for each of 20 consecutive simulation runs per scenario. Prevalence was not separated into prevalence among the vaccinated or unvaccinated but was rather the total prevalence in the 40 agents.

**Table 1:** Scenario settings varied included vaccination rate, the masking status of the single initial infectious agent, and the masking status of the 39 initially susceptible agents.

| ID | Scenario | Vaccination Rate | Infectious Unmasked | Infectious Masked | Susceptible Unvaccinated Unmasked | Susceptible Unvaccinated Masked | Susceptible Vaccinated Unmasked | Susceptible Vaccinated Masked |
| --- | --- | --- | --- | --- | --- | --- | --- | --- |
| A | Half vaccination, infectious unmasked, susceptibles unmasked | 52.5 | 1 | 0 | 18 | 0 | 21 | 0 |
| B | Half vaccination, infectious unmasked, susceptibles half masked | 52.5 | 1 | 0 | 9 | 9 | 10 | 11 |
| C | Half vaccination, infectious unmasked, susceptibles all masked | 52.5 | 1 | 0 | 0 | 18 | 0 | 21 |
| D | Half vaccination, infectious masked, susceptibles unmasked | 52.5 | 0 | 1 | 18 | 0 | 21 | 0 |
| E | Half vaccination,<br>infectious masked,<br>susceptibles half masked | 52.5 | 0 | 1 | 9 | 9 | 10 | 11 |
| F | Half vaccination,<br>infectious masked,<br>susceptibles all masked | 52.5 | 0 | 1 | 0 | 18 | 0 | 21 |
| G | 100% vaccination,<br>infectious unmasked,<br>susceptibles unmasked | 100 | 1 | 0 | 0 | 0 | 39 | 0 |
| H | 100% vaccination,<br>infectious unmasked,<br>susceptibles half masked | 100 | 1 | 0 | 0 | 0 | 19 | 20 |
| I | 100% vaccination,<br>infectious unmasked,<br>susceptibles all masked | 100 | 1 | 0 | 0 | 0 | 0 | 39 |
| J | 100% vaccination,<br>infectious masked,<br>susceptibles unmasked | 100 | 0 | 1 | 0 | 0 | 39 | 0 |
| K | 100% vaccination,<br>infectious masked,<br>susceptibles half masked | 100 | 0 | 1 | 0 | 0 | 19 | 20 |
| L | 100% vaccination,<br>infectious masked,<br>susceptibles all masked | 100 | 0 | 1 | 0 | 0 | 0 | 39 |

The infectious agent either was or was not masked. Susceptible agents were either vaccinated or unvaccinated according to the percentage vaccination being tested. Masking in the susceptible agents was none, half, or all, with half representing partial masking of the vaccinated (and partial masking of the unvaccinated in the 52% vaccination scenarios). Although the partial masking scenario is referred to as “half,” due to there being 39 agents, 21 of whom were vaccinated in the 52% vaccination scenarios, this was not always exactly half as agents in a group cannot be a decimal value. The larger number was given to the masked group in each scenario’s settings. The exception to this is the unvaccinated in the 52% trial runs, as there were 18 in this group.

The quanta rate was taken from a paper estimating it from an early outbreak on a bus in China as 0.238 per second, or 14.28 per minute [9]. To model the Delta variant, the quanta of which is not yet known, reports that R_0_ for Delta may be approximately four times higher than for the original strain were extrapolated to multiply the original quanta by four to get 57.12 quanta per minute [10]. Vaccine efficacy was set at 80%, as a study showed that this was the approximate efficacy against asymptomatic infection, which is sufficient for transmission, with the original strain after vaccination with the BioNTech/Pfizer vaccine [11]. It was assumed that efficacy against Delta is no better than against the original strain, so this represents a conservative best-case scenario. Mask efficacy was set at 95% to model the usage of N95 masks, which, while not common, represents a best-case conservative efficacy.

Poisson regression was used for statistical analysis in R, with prevalence at the end of the simulation as the response variable, and predictor variables of whether the infectious agent was masked or not, whether the susceptible agent group was 52% vaccinated (called “half”) or 100% vaccinated (called “all”), and whether masking of the susceptible agents was classed as none, half, or all.

## Results

The significance of predictor variables in determining prevalence was tested using an ANOVA approach on the Poisson regression (Table 2). Scenarios with the infectious agent masked had a significantly lower prevalence than the exact same scenario in which the infectious agent was not masked, regardless of other factors (p < 2.2E-16, Figure 1 and Table 2, see Appendix for data). The highest prevalence was seen, in both vaccination scenarios, when neither the infectious agent nor any of the susceptible agents were masked (scenarios A and G), and varying masking of the susceptibles also showed a highly significant effect on prevalence (P < 2.2E-16). There was not a significant difference between 52% vaccination and 100% vaccination in general (p = 0.326162). There was a highly significant interaction between the masking of susceptibles and the masking of the infectious agent (p < 1.38E-05). The interaction between masking and vaccination was also significant (p < 0.024), and the interaction between all three variables was also significant (p < 0.0062). However, the interaction between vaccination and the masking of the infectious agent was not significant (p = 0.390809). The lowest prevalence was seen when 100% were vaccinated, the infectious agent was masked, and all susceptible agents were masked (scenario L). This was significantly lower than the same masking scenario with 52% (“half”) vaccination (scenario F).

**Table 2:** ANOVA table for the Poisson regression with Prevalence as the response variable and Infectious_masked, Masking, and Vaccination as the predictor variables.

| Response: Prevalence |  |  |  |  |  |  |  |  |  |  |  |  |
| --- | --- | --- | --- | --- | --- | --- | --- | --- | --- | --- | --- | --- |
|  | LR Chi sq | D f | Pr (> Chi sq) | Significance |  |  |  |  |  |  |  |  |
| Infectious_masked | 345.68 | 1 | < 2.2E-16 | *** |  |  |  |  |  |  |  |  |
| Masking | 140.93 | 2 | < 2.2E-26 | *** |  |  |  |  |  |  |  |  |
| Vaccination | 0.96 | 1 | 0.326162 |  |  |  |  |  |  |  |  |  |
| Infectious_masked:Masking | 22.38 | 2 | 1.38E-05 | *** |  |  |  |  |  |  |  |  |
| Infectious_masked:Vaccination | 0.74 | 1 | 0.390809 |  |  |  |  |  |  |  |  |  |
| Masking:Vaccination | 7.51 | 2 | 0.023375 | * |  |  |  |  |  |  |  |  |
| Infectious_masked:Masking:Vaccination | 10.17 | 2 | 0.006176 | ** |  |  |  |  |  |  |  |  |
| Significance codes: |  | 0 | '***' | 0.001 | '**' | 0.01 | '**' | 0.05 | '.' | 0.1 | ' ' | 1 |

**Figure 1:**
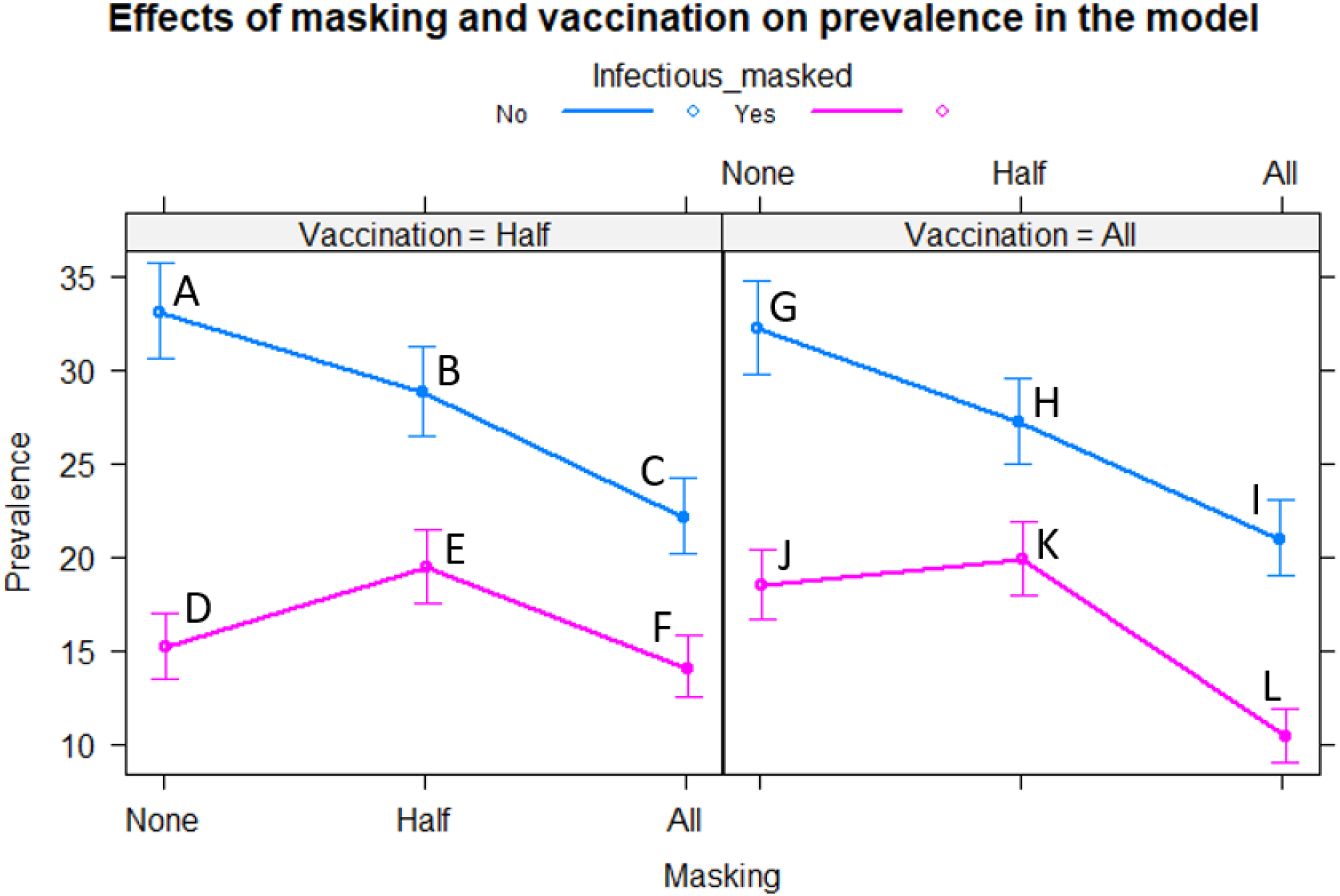
The effects plot showing the prevalence as affected by vaccination, masking of the infectious agent, and masking of the susceptible agents. Letter labels refer to the scenarios in Table 1. Error bars show the 95% confidence interval.

## Discussion

The emergence of the Delta variant has changed the landscape of the response to COVID-19. If people are to mix as before in public places (which is the desired outcome), the best way to protect both people and the ability to conduct usual business is to reduce the spread of this dangerous variant, which is now the dominant variant in the U.S as of the time of writing [12]. Although vaccination is doubtlessly important (especially in reducing morbidity and mortality, which it does very well) and, as a society, we should continue to support as many getting vaccinated as possible, vaccination alone is not enough to ensure a reasonably safe level of exposure during usual business, such as returning to in-person classes in university settings. Some people, including all children under 12 currently, will not have a choice whether to get vaccinated, and medical and religious exemptions, as well as incomplete immunity in vaccinated but immunocompromised individuals, means even very high rates of vaccination do not preclude the existence of vulnerable subpopulations. Additionally, Delta is too infectious and asymptomatic cases are too likely to rely on vaccination alone, which is, *at most*, 80% effective against infection with the Delta variant (based on data on previous strains, [11]) and may be as low as 66% [13]. Therefore, masking becomes an important consideration for reducing transmission of COVID-19 as society continues to live in the reality of an ongoing pandemic.

Although vaccination was not significant in the analysis, that is not to say it is not important; vaccination greatly reduces disease severity, hospitalization, and death from COVID-19. However, it is less effective against infection, including mild or asymptomatic infection. These results show that, regardless of vaccination (even in the context of the ideal 100% vaccination), masking is an important barrier to the spread of COVID-19. Masking even half of the susceptible population reduces prevalence in these scenarios, but the most important determinant is whether the infectious agent is masked. The lowest infection, with realistic (52%) or ideal vaccination (100%), is obtained from all agents masking: the infectious agent and all of the susceptible agents.

An important strength of COVID-ADAPT is the ability to model asymptomatic infections. In this model, it is irrelevant whether the initial infectious agent is symptomatic: regardless, it is capable of infecting others. Considering that the initially infectious agent may be taken to possibly be asymptomatic, the importance of masking mandates becomes clear. The best results are when everyone is masked, but even if some compliance slips and a few (such as the initially infectious agent) are not adequately masked, the likelihood of at least some of those noncompliant people being asymptomatically infected makes infection of others highly likely. The best way to greatly reduce infection, regardless of vaccination status, is to ensure that infectious people are masked in addition to susceptible people. And due to the possibility of asymptomatic infection (estimated to be 30% of COVID-19 infections as of writing), the only way to mask all the infectious individuals is to have universal mask mandates and ensure compliance [14].

Like any model that seeks to simulate aspects of reality while remaining tractably simplified, COVID-ADAPT has limitations. Chief among these is that the behavior of infected agents does not change, such as with isolation or even reduced movement if the individual were symptomatic. However, these scenarios are modeling a dorm floor in which, even if an individual is isolating in their room, might still use restrooms and common hallways. In previous terms, students testing positive at EWU were removed to quarantine dorms to reduce this sort of incidental exposure to others. Undoubtedly, though, this only concerned students tested, whether by choice or due to contact tracing turning up exposure. Therefore, asymptomatically infected students who were not aware of their exposure, occurring perhaps in a semi-anonymous public space, such as the grocery store or the cafeteria, where contact tracing is unlikely, would still go about their business on their dorm floors, potentially infecting others.

In this way, the asymptomatic possibility provides reasoning for this scenario. Additionally, COVID-ADAPT as a model is intended to provide data on whether specific interventions reduce spread, and this is currently best modeled with a contained population. For this reason, too, death, hospitalization, and other causes of removal from the floor are not modeled. Future iterations of the model may include more complex movement behaviors, including movement to other floors and eventually other buildings, and at such time, quarantining may be a desirable outcome of infection in the model. This too answers concerns that the behavior of the agents, being randomized and confined to one floor for three weeks, is unrealistic: as a simplified model, it serves its purpose, and later development will introduce behavioral rules for movement, perhaps even to include class schedules and other movement between buildings. Also, one could consider the simulation to represent not three continuous weeks, but three weeks’ worth of exposure within a dorm floor of the residents there. In this communal housing scenario, large amounts of overlap between students’ presence in the building makes this a reasonable avenue of exposure.

One additional concern is that N95 masks were modeled. These are not common, nor are they required in mask mandates for any but certain medical personnel in high-risk environments. Certainly, mask efficacy affects the overall protective effect of masking, and trials with less efficacious masks such as surgical masks or the popular cloth masks would likely result in different numbers. However, using N95 masks as the standard for this study represents a conservative estimate of how the best-case scenario of masking would affect transmission in such a situation. Because of this, it is likely the change would be in the degree of protection, not the overall trend. Since vaccination does not provide sterilizing immunity, allowing asymptomatic infection in addition to breakthrough symptomatic infections, it is reasonable to test what other measures in addition would protect the population the best. Real-world mask efficacy is difficult to estimate as a wide variety of materials and qualities are used, variations in mask compliance and hygiene exist, and the effects of improperly fitting or improperly worn masks mitigate their efficacy. For the purposes of modeling the effect of masks, N95s are a good starting point, and future studies utilizing different mask efficacies are certainly warranted.

Finally, the difficulty of establishing parameters must be addressed. For as much as it feels like the reign of COVID-19 has been interminable, in terms of human diseases, SARS-CoV-2 is in its infancy. Little is known about it, and much of what is known is back-calculated (with all of the attendant margin for error) from specific events or is tentatively estimated. There has not been enough time to establish with strong certainty parameters such as viral decay, vaccine efficacy against variants, and quanta emitted. Quanta emitted by vaccinated persons may likely turn out to be lower than that emitted by unvaccinated persons. Additionally, the quick establishment of Delta’s dominance in many places has changed the degree of reliability of earlier statistics and experiments. Nonetheless, for COVID-ADAPT, parameters were either sourced from the best literature available at this point (such as for vaccine efficacy and quanta) or chosen to establish a consistent baseline against which to test other measures (such as for mask efficacy and viral decay). Continued research on COVID-19 in the greater scientific community is being doggedly pursued, and no doubt as time goes on, data with greater certainty will emerge and can then be incorporated into this flexible model.

As a preliminary test of COVID-ADAPT, the results show the model performed according to reasonable expectations. Further development of the model is planned, with changes such as easier custom map creation to model the building(s) of most concern to the user, behavioral movement rules or even scheduled behaviors for agents (such as class or sleep schedules), and improvements to visual output on the list of desired features. Additionally, the emergence of more and better data will allow greater pinpointing of parameters for particular scenarios. The data from these trials, however, show that, regardless of the differences between complex real-life situations and streamlined models, universal masking is important in reducing transmission of COVID-19. Vaccination alone, even at the idealized 100% uptake, does not sufficiently mitigate the spread of highly infectious strains like Delta, particularly when asymptomatic infections are concerned.

COVID-19 is a test of collective willpower and empathy as we, as a community, country, and global society, struggle to balance established health interventions such as vaccination and masking with the miasma of uncertainty and misinformation that clouds the issue. The availability of more and better data is itself a welcome “vaccination” against this uncertainty, and this is the niche that COVID-ADAPT fills: even in a simple, early build, this model and these trials show that the collection and analysis of data through multiple avenues (including simulation models) will help support decision-making about health interventions. Masking in particular is shown to be a vital measure to adopt in the time of the Delta variant as society continues to open up. With sound data and well-reasoned policy decisions, steps can be taken to mitigate the effect that COVID-19 has on the health and welfare of communities and the individuals in care of those communities. The data show that universal masking mandates are a vital application of this principle. By reducing transmission by even the asymptomatically infectious and protecting the susceptible, masking, in addition to the continued campaign to vaccinate as many as possible, can significantly reduce the consequences of COVID-19 in communities such as universities.

## Data Availability

All data is included in the manuscript.

## Appendix: Data

**Table 3:**
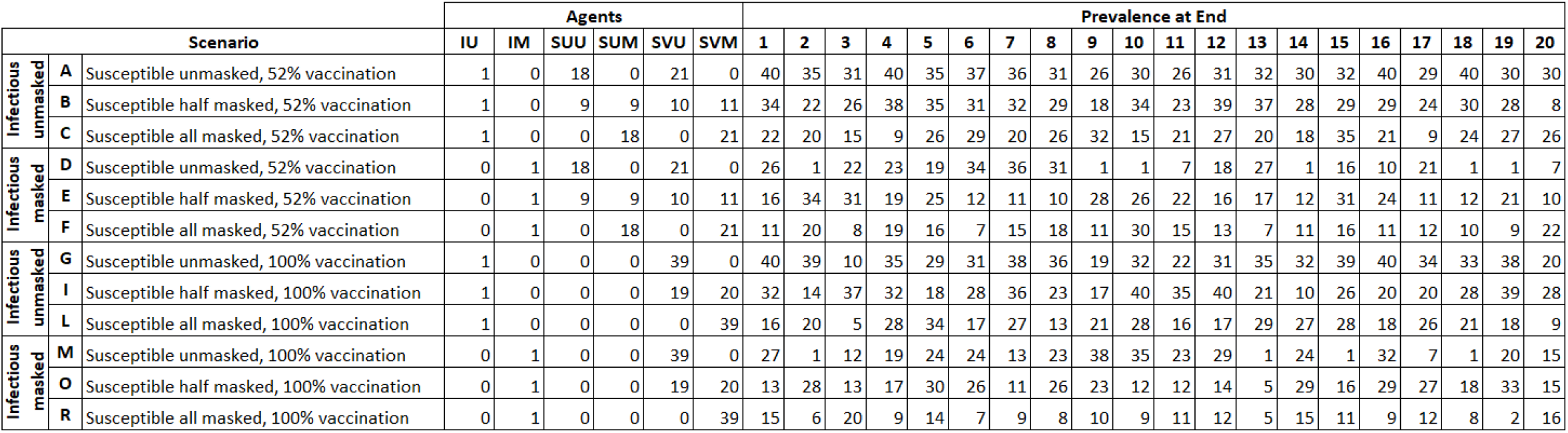
The 12 scenarios, divided by masking of the infectious agent and susceptible agents and vaccination percentage, were run 20 times each. The prevalence at the end of the simulation includes the initial infectious person. Abbreviations: IU – infectious agent (unmasked), IM – infectious agent (masked), SUU – susceptible agents (unvaccinated, unmasked), SUM – susceptible agents (unvaccinated, masked), SVU – susceptible agents (vaccinated, unmasked), and SVM susceptible agents (vaccinated, masked).

